# Progression of Frailty and Cardiovascular Outcomes Among Medicare Beneficiaries

**DOI:** 10.1101/2024.02.09.24302612

**Authors:** Yusi Gong, Yang Song, Jiaman Xu, Huaying Dong, Ariela R. Orkaby, Daniel B. Kramer, John A. Dodson, Jordan B. Strom

## Abstract

**Background:** Frailty is associated with adverse cardiovascular outcomes independent of age and comorbidities, yet the independent influence of frailty progression remains uncertain.

**Methods:** Medicare Fee-for-service beneficiaries ≥ 65 years at cohort inception with continuous enrollment from 2003-2015 were included. Frailty trajectory was measured by annualized change in a validated claims-based frailty index (CFI) over a 5-year period. Linear mixed effects models, adjusting for baseline frailty, were used to estimate CFI change over a 5-year period. Survival analysis was used to evaluate associations of frailty progression and future health outcomes (major adverse cardiovascular and cerebrovascular events [MACCE], all-cause death, heart failure, myocardial infarction, ischemic stroke, and days alive at home [DAH] within the following calendar year).

**Results:** 26.4 million unique beneficiaries were included (mean age 75.4 ± 7.0 years, 57% female, 13% non-White). In total, 20% had frailty progression, 66% had no change in frailty, and 14% frailty regression over median follow-up of 2.4 years. Compared to those without a change in CFI, when adjusting for baseline frailty, those with frailty progression had significantly greater risk of incident MACCE (hazard ratio [HR] 2.30, 95% confidence interval [CI] 2.30-2.31), all-cause mortality (HR 1.59, 95% CI 1.58-1.59), acute myocardial infarction (HR 1.78, 95% CI 1.77-1.79), heart failure (HR 2.78, 95% CI 2.77-2.79), and stroke (HR 1.78, 95% CI 1.77-1.79). There was also a graded increase in risk of each outcome with more rapid progression and significantly fewer DAH with the most rapid vs. the slowest progression group (270.4 ± 112.3 vs. 308.6 ± 93.0 days, rate ratio 0.88, 95% CI 0.87-0.88, p < 0.001).

**Conclusions:** In this large, nationwide sample of Medicare beneficiaries, frailty progression, independent of baseline frailty, was associated with fewer DAH and a graded risk of MACCE, all-cause mortality, myocardial infarction, heart failure, and stroke compared to those without progression.

**NON-STANDARD ABBREVIATIONS AND ACRONYMS:**

- Claims-based frailty index (CFI)
- Major adverse cardiovascular and cerebrovascular events (MACCE)
- Number of days alive at home within the following calendar year (DAH)
- Medicare Fee-for-Service (FFS)

## INTRODUCTION

Frailty, a state of decreased physiologic reserve resulting in diminished capacity to maintain homeostasis after a stressor, predicts the risk of adverse health outcomes better than chronologic age and burden of comorbidities.^1–6^ Multiple validated tools have been developed for measuring frailty,^6–9^ with the goal of improving risk stratification and medical decision-making in a variety of settings including treatment strategies for aortic stenosis,^10–12^ anticoagulation for atrial fibrillation,^13–15^ and cardiac resynchronization therapy.^16^ However, as most frailty studies are cross-sectional, few analyses have evaluated the natural history of frailty in older adults in the United States^17–19^ and even fewer have focused specifically on its relation to cardiovascular outcomes.^20^ Understanding frailty trajectories may be important given the growing focus on pre-rehabilitation interventions designed to modify frailty before high-risk cardiovascular procedures.^21,22^

Medicare claims represent an attractive setting to evaluate the natural history of frailty. As frailty ultimately manifests itself through contacts with the healthcare system, the digital signature of these encounters, namely billing claims, can be used to identify frail individuals and assess how their burden of frailty changes over time. Claims-based frailty indices (CFIs) have been developed against the reference standard of in-person frailty measurements and shown to identify impairments in function and risk for adverse outcomes similar to in-person measures.^8,9,11,23^ However, data are limited on how changes in CFI are associated with risk of adverse cardiovascular events. As such, we sought to leverage complete Medicare claims to estimate expected changes in CFI over time and evaluate if rate of frailty progression independently identifies individuals at higher risk of adverse cardiovascular outcomes.

## METHODS

### Data Source

We analyzed data from the United States Centers for Medicare and Medicaid Services (CMS) Medicare Provider Analysis and Review (MedPAR) database from January 1, 2003 through December 31, 2020. The MedPAR database consists of a 100% sample of inpatient discharge claims and services (Part A claims) for Medicare Fee-for-Service (FFS) beneficiaries and has been used extensively in evaluation of health policy and practice.^24–26^

### Study Population

As Medicare-eligible individuals may include those on disability or needing renal replacement therapy, individuals < 65 years old on cohort entry were excluded. Additionally, to define a cohort with continuous enrollment in which frailty trajectory could be reliably assessed, we excluded individuals without at least 5 years of continuous enrollment in the Medicare FFS program from January 1, 2003 to September 30, 2015 (i.e. the exposure period). The study was approved by the Beth Israel Deaconess Medical Center Institutional Review Board with a waiver of informed consent. The data used in this analysis are not available for public sharing due to data use agreements with Medicare (RSCH-2018-52411).

### Covariates

Patient-level covariates included age on cohort entry, sex, self-reported race and ethnicity, and 27 Medicare Chronic Condition Data Warehouse indicators which are derived from validated algorithms for relevant comorbidities based on a combination of inpatient and outpatient codes in preceding 2 years.^27^

### Definition of Exposure

Frailty was defined using the Johns-Hopkins CFI (**Supplemental Table 1**).^8^ This CFI was derived based on the gold-standard for in-person frailty assessment, the Fried frailty phenotype,^6^ which itself was derived from the Cardiovascular Health Study and validated in the National Health and Aging Trends study.^28^ We have previously demonstrated that this CFI identifies individuals with greater impairment in functional outcomes such as activities of daily living (ADLs) and predicts outcomes similar to in-person metrics of frailty.^10,29^ To determine frailty trajectory, we generated this CFI for each individual in the cohort during the first five years of enrollment in the exposure period between January 1, 2003 and September 30, 2015. We used the linear trend in CFI across these five years to determine frailty trajectory slope (see “Statistical Methods” below). Baseline frailty was assessed at the end of the exposure period using a CFI ≥ 0.2 to define frailty.^23^

### Outcomes

The primary endpoint was major adverse cardiovascular and cerebrovascular events (MACCE), defined as the composite of total death, acute myocardial infarction (MI), ischemic stroke, or heart failure (HF) hospitalization. Secondary endpoints were all-cause mortality, incident HF, MI, or ischemic stroke, ascertained using vital status information in the linked Medicare Beneficiary Summary File. Outcomes were defined using claims algorithms (**Supplemental Table 2**) previously validated against trial-adjudicated endpoints, using the first occurrence of a claim to define an event. Additionally, we evaluated the number of days alive at home (DAH) within one-year as a secondary endpoint, excluding hospitalizations and rehabilitation stays as per prior methods.^30,31^ Death information was complete for all individuals. Individuals were followed for the occurrence of an event from the end of the 5-year exposure period through December 31, 2020.

### Statistical Analysis

Linear mixed effects models with a random intercept and slope for time since the start of cohort entry were used to estimate frailty trajectory over the 5-year exposure period, adjusting for baseline CFI. Specifically, we estimated the conditional predicted slope for time (i.e. each individual’s predicted change in CFI over the 5-year period) by adding the estimated individual random effects for time to the marginal slope for time. Based on these results, the cohort was split into three frailty trajectory groups according to change in slope of CFI over the exposure period: conditional predicted slope = 0.09 (unchanged frailty; representing individuals without hospitalizations for whom the only change in CFI over the 5-year exposure period was an increase in age), conditional predicted slope < 0.09 (frailty regressors; representing regression in CFI over the 5-year period), and conditional predicted slope > 0.09 (frailty progressors; representing progression in CFI over the 5-year period). Baseline characteristics at the start of follow-up were determined, stratified across the three frailty trajectory groups, using means and standard deviations (SD) or medians and interquartile ranges (IQR) for continuous variables and numbers and proportions for categorical variables.

The group with frailty progression was subsequently further split into quartiles of CFI progression. Baseline characteristics of those with frailty progression were determined, stratified by progression quartile, similar to above and compared across groups using the absolute value of standardized mean differences to assess for imbalance, with a value ≥ 0.1 considered a significant imbalance.^32–34^ Cumulative incidence functions were generated for time from the end of the exposure period to the first occurrence of each outcome, stratified by frailty trajectory (regressors, unchanged frailty, and progressors) and quartiles of frailty progression, comparing across groups using log-rank tests. Using Cox proportional hazards models, the hazard ratio (HR) and 95% confidence interval (CI) for time to occurrence of each outcome were determined by frailty trajectory and quartiles of frailty progression and 5-year outcome rates estimated. Alive individuals were censored at the end of follow-up. Negative binomial regression was used to estimate the rate ratio for DAH by frailty trajectory and quartiles of frailty progression. All analyses were conducted using SAS v9.4 (SAS Institute, Cary, NC) using a two-tailed p-value < 0.05 for significance.

## RESULTS

### Overall Cohort

During the exposure period, there were 83,023,168 unique individuals enrolled in the Medicare FFS program. Of these, 16,299,009 (19.6%) were excluded for age < 65 years, 30,500,369 (36.7%) for non-continuous enrollment, and 9,808,903 (11.8%) for not having follow-up between 2009-2020, before death, or before last consecutive FFS coverage date. A total of 26,414,887 beneficiaries met inclusion for the final cohort (**Figure 1**; mean age 75.4 ± 7.0, 57% female, 87% White). A total of 66.1% of the cohort had no change in frailty, 13.6% had frailty regression, and 20.4% had frailty progression. A total of 7.3% were considered frail (CFI ≥ 0.2) at baseline, including 21.8% of frailty progressors, 7.2% of frailty regressors, and 2.9% of the unchanged frailty group (**Table 1**).

**Figure 1.**
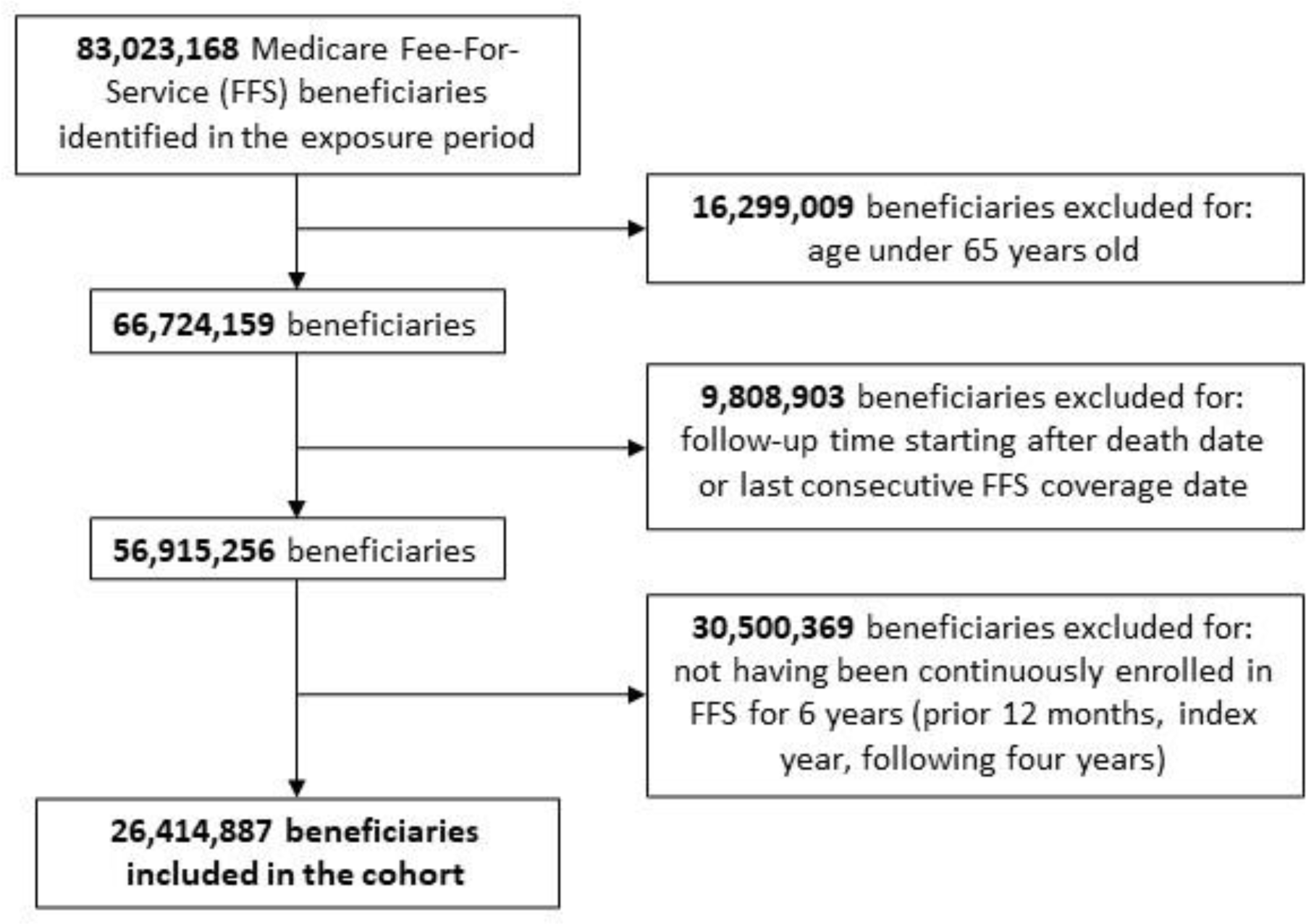
Flow Diagram Illustrating Study Inclusion and Exclusion Criteria. Flow diagram for inclusion and exclusion of beneficiaries. Exclusion criteria were: (1) beneficiaries < 65 years old, (2) beneficiaries who did not have appropriate follow-up (eg. died before follow-up, did not have coverage throughout follow-up, or did not have follow-up in years 2009-2015), and (3) beneficiaries without at least 6 years of continuous enrollment (5 years needed to define a baseline frailty trajectory plus at least one year of follow-up).

**Table 1.** Baseline Characteristics at the Start of Follow-up. ^a^Calculated using a claims-based frailty index value ≥ 0.2. ^b^Represents the standardized mean difference (SMD) between the frailty regressor group (claims-based frailty index slope > 0.09) vs. unchanged frailty group (claims-based frailty index slope = 0.09). ^c^Represents the SMD between the frailty progressor group (claims-based frailty index slope > 0.09) vs. unchanged frailty group. Baseline characteristics were estimated at the end of the 5-year exposure period (i.e. the start of event follow-up). A SMD ≥ 0.1 was considered evidence of significant group imbalance and is indicated in bold font. Numbers are listed as percentages unless otherwise indicated. SD = standard deviation, Q1 = quartile 1, Q3 = quartile 3.

| Subject Characteristic | Overall<br>(N =<br>26,274,544) | Frailty<br>Regressors<br>(N =<br>3,563,528) | Unchanged<br>Frailty<br>(N =<br>17,354,604) | Frailty<br>Progressors<br>(N =<br>5,356,412) | SMD for<br>Frailty<br>(Regressors vs.<br>Unchanged<br>Frailty)<sup>b</sup> | SMD for<br>Frailty<br>(Progressors<br>vs. Unchanged<br>Frailty)<sup>c</sup> |
| --- | --- | --- | --- | --- | --- | --- |
| Baseline Frailty Present<sup>a</sup> | 7.3% | 7.2% | 2.9% | 21.8% | **0.20** | **0.60** |
| Age |  |  |  |  |  |  |
| Mean ± SD | 75.4±7.0 | 77.4±7.2 | 74.4±6.7 | 77.2±7.3 | **0.43** | **0.40** |
| Median (Q1, Q3) | 73.0<br>(69.0,80.0) | 77.0<br>(71.0,83.0) | 72.0<br>(69.0,78.0) | 76.0<br>(70.0,83.0) |  |  |
| (Min, Max) | (69.0,119.0) | (69.0,119.0) | (69.0,119.0) | (69.0,119.0) |  |  |
| Male | 43.0% | 41.3% | 43.9% | 41.3% | 0.05 | 0.05 |
| Race |  |  |  |  |  |  |
| White | 87.0% | 89.4% | 85.9% | 88.9% | **0.11** | 0.09 |
| Black | 7.0% | 6.6% | 7.0% | 6.9% | 0.02 | **0.40** |
| Other | 6.0% | 3.9% | 7.1% | 4.2% | **0.14** | **0.13** |
| Prior Myocardial Infarction | 3.5% | 8.4% | 1.0% | 8.5% | **0.35** | **0.36** |
| Alzheimer's Disease | 4.4% | 7.5% | 2.3% | 8.9% | **0.24** | **0.29** |
| Alzheimer's Disease and Related Disorders or Senile Dementia | 9.5% | 16.1% | 5.2% | 19.1% | **0.36** | **0.44** |
| Atrial Fibrillation | 11.3% | 20.9% | 5.9% | 22.7% | **0.45** | **0.50** |
| Cataract | 59.9% | 71.2% | 54.6% | 69.4% | **0.35** | **0.31** |
| Chronic Kidney Disease | 12.8% | 21.8% | 6.4% | 27.3% | **0.45** | **0.58** |
| Chronic Obstructive Pulmonary Disease | 19.8% | 33.6% | 12.0% | 35.7% | **0.53** | **0.58** |
| Heart Failure | 20.8% | 38.6% | 10.9% | 41.1% | **0.68** | **0.73** |
| Diabetes | 28.0% | 38.0% | 22.5% | 39.2% | **0.34** | **0.37** |
| Glaucoma | 19.1% | 22.2% | 17.5% | 21.9% | **0.12** | **0.11** |
| Hip/Pelvic Fracture | 2.7% | 5.9% | 0.7% | 6.9% | **0.30** | **0.33** |
| Ischemic Heart Disease | 41.4% | 65.0% | 29.3% | 64.6% | **0.77** | **0.76** |
| Depression | 19.3% | 30.4% | 13.2% | 31.7% | **0.43** | **0.45** |
| Osteoporosis | 17.8% | 24.3% | 14.3% | 24.7% | **0.25** | **0.26** |
| Rheumatoid Arthritis / Osteoarthritis | 43.8% | 62.6% | 34.2% | 62.4% | **0.59** | **0.59** |
| Stroke / Transient Ischemic Attack | 11.4% | 21.8% | 5.8% | 22.7% | **0.48** | **0.50** |
| Cancer | 13.3% | 19.9% | 10.0% | 19.5% | **0.28** | **0.27** |
| Anemia | 41.0% | 62.5% | 29.2% | 64.7% | **0.71** | **0.76** |
| Baseline Frailty Present<sup>a</sup> | 7.3% | 7.2% | 2.9% | 21.8% | **0.20** | **0.60** |
| Asthma | 9.0% | 14.7% | 5.9% | 15.1% | **0.29** | **0.31** |
| Hyperlipidemia | 66.1% | 78.4% | 59.8% | 78.2% | **0.42** | **0.41** |
| Benign Prostatic<br>Hyperplasia | 15.3% | 20.0% | 12.8% | 20.3% | **0.20** | **0.20** |
| Hypertension | 71.3% | 88.8% | 62.0% | 89.7% | **0.66** | **0.69** |
| Acquired Hypothyroidism | 20.0% | 26.8% | 16.5% | 26.7% | **0.25** | **0.25** |

The mean annual change in CFI was 0.10 ± 0.09 overall, 0.21 ± 0.11 for frailty progressors, and 0.001 ± 0.09 for frailty regressors. The mean annual change in CFI was numerically similar between males (0.10 ± 0.09) and females (0.10 ± 0.08). Similarly, the mean annual change in CFI was numerically similar between Whites (0.10 ± 0.09), Blacks (0.10 ± 0.09), and other races (0.10 ± 0.07). Those with unchanged frailty were overall younger and had lower prevalence of all baseline comorbidities (**Table 1**).

### Characteristics of Frailty Progressors

Among frailty progressors, 26% were in quartile 1 (mean annual change in CFI 0.11 ± 0.01, range 0.09-0.13), 24% in quartile 2 (mean annual change in CFI 0.15 ± 0.01, range 0.13-0.17), 25% in quartile 3 (mean annual change in CFI 0.21 ± 0.02, range 0.17-0.25), and 25% in quartile 4 (mean annual change in CFI 0.36 ± 0.11, range 0.25-1.33) (**Supplemental Figure 1**). With increasing quartiles of frailty progression, there was increasing prevalence of all comorbidities (**Table 2**).

**Table 2.** Baseline Characteristics Stratified by Frailty Progression Quartile. ^a^Calculated using a claims-based frailty index value ≥ 0.2. ^b^Represents the standardized mean difference (SMD) between quartile 4 (Q4) of frailty progressors (fastest progression) vs. quartile 1 (Q1) of frailty progressors (slowest progression). Frailty progression was defined using a claims-based frailty index slope > 0.09. A SMD ≥ 0.1 was considered evidence of significant group imbalance and is indicated in bold font. Numbers are percentages unless otherwise indicated. SD = standard deviation, Q1 = quartile 1, Q3 = quartile 3.

| Subject Characteristic | Overall<br>(N =<br>5,356,412) | Quartile 1<br>(N =<br>1,394,145) | Quartile 2<br>(N =<br>1,294,208) | Quartile 3<br>(N =<br>1,351,532) | Quartile 4<br>(N =<br>1,316,527) | SMD for<br>Frailty<br>(Q4 vs.<br>Q1)<sup>b</sup> |
| --- | --- | --- | --- | --- | --- | --- |
| Baseline Frailty Present<sup>a</sup> | 21.8% | 6.1% | 9.1% | 18.9% | 53.7% | **1.22** |
| Age |  |  |  |  |  |  |
| Mean ± SD | 77.2±7.3 | 76.1±6.9 | 76.4±7.0 | 77.4±7.3 | 78.9±7.6 | **0.39** |
| Median (Q1, Q3) | 76.0<br>(70.0,83.0) | 75.0<br>(69.0,81.0) | 75.0<br>(69.0,81.0) | 76.0<br>(70.0,83.0) | 79.0<br>(72.0,85.0) |  |
| (Min, Max) | (69.0,119.0) | (69.0,112.0) | (69.0,119.0) | (69.0,113.0) | (69.0,111.0) |  |
| Male | 41.3% | 44.4% | 44.6% | 39.8% | 36.2% | **0.17** |
| Race |  |  |  |  |  |  |
| White | 88.9% | 89.1% | 89.2% | 89.0% | 88.1% | 0.03 |
| Black | 6.9% | 6.3% | 6.6% | 7.0% | 8.0% | 0.07 |
| Other | 4.2% | 4.6% | 4.2% | 4.0% | 3.9% | 0.03 |
| Prior Myocardial Infarction | 8.5% | 6.4% | 8.3% | 8.3% | 11.2% | **0.17** |
| Alzheimer's Disease | 8.9% | 3.7% | 6.0% | 9.3% | 16.8% | **0.44** |
| Alzheimer's Disease and Related Disorders or Senile Dementia | 19.1% | 9.5% | 13.1% | 19.7% | 34.6% | **0.64** |
| Atrial Fibrillation | 22.7% | 18.2% | 18.9% | 23.2% | 30.7% | **0.29** |
| Cataract | 69.4% | 67.5% | 67.8% | 69.7% | 72.5% | **0.11** |
| Chronic Kidney Disease | 27.3% | 20.2% | 23.9% | 27.7% | 38.0% | **0.40** |
| Chronic Obstructive Pulmonary Disease | 35.7% | 26.2% | 33.6% | 36.9% | 46.7% | **0.43** |
| Heart Failure | 41.1% | 26.6% | 31.6% | 43.3% | 63.3% | **0.79** |
| Diabetes | 39.2% | 35.3% | 36.5% | 39.3% | 45.8% | **0.22** |
| Glaucoma | 21.9% | 21.4% | 21.2% | 21.8% | 23.0% | 0.04 |
| Hip/Pelvic Fracture | 6.9% | 4.9% | 4.8% | 6.8% | 11.3% | 0.24 |
| Ischemic Heart Disease | 64.6% | 59.6% | 61.4% | 64.0% | 73.7% | **0.30** |
| Depression | 31.7% | 20.3% | 23.9% | 33.4% | 49.9% | **0.65** |
| Osteoporosis | 24.7% | 22.1% | 21.4% | 25.3% | 30.1% | **0.18** |
| Rheumatoid Arthritis / Osteoarthritis | 62.4% | 52.0% | 59.6% | 66.9% | 71.7% | **0.41** |
| Stroke / Transient Ischemic Attack | 22.7% | 14.5% | 19.3% | 24.2% | 33.1% | **0.45** |
| Cancer | 19.5% | 19.6% | 21.3% | 18.6% | 18.6% | 0.03 |
| Anemia | 64.7% | 55.4% | 60.7% | 66.8% | 76.4% | **0.45** |
| Asthma | 15.1% | 11.2% | 14.7% | 15.6% | 19.2% | **0.22** |
| Hyperlipidemia | 78.2% | 78.3% | 78.3% | 77.9% | 78.1% | 0.06 |
| Benign Prostatic Hyperplasia | 20.3% | 21.5% | 21.0% | 19.5% | 19.1% | 0.06 |
| Hypertension | 89.7% | 86.6% | 88.2% | 90.3% | 94.0% | **0.25** |
| Acquired Hypothyroidism | 26.7% | 24.5% | 24.7% | 27.1% | 30.8% | **0.14** |

### Outcomes of Frailty Progressors vs. Regressors

Over a median follow-up of 2.4 years, 48.6% experienced MACCE (vs. 22.8% with unchanged frailty; HR, 95% CI: 2.30, 2.30-2.31) (**Figure 2**). 33.7% of those with frailty progression died (vs. 16.1% with unchanged frailty; HR, 95% CI: 2.32, 2.32-2.33). A total of 5.7% of those with frailty progression had acute MI (vs. 3.0% with unchanged frailty; HR, 95% CI: 1.78, 1.77-1.79), 12.1% experienced HF (vs. 4.0% with unchanged frailty; HR, 95% CI: 2.78, 2.77-2.79), and 10.2% experienced an ischemic stroke (vs. 5.3% with unchanged frailty; HR, 95% CI: 1.78, 1.77-1.79) (**Table 3**). Those with frailty regression had overall worse outcomes compared to the unchanged frailty group, but better outcomes than those with frailty progression.

**Figure 2.**
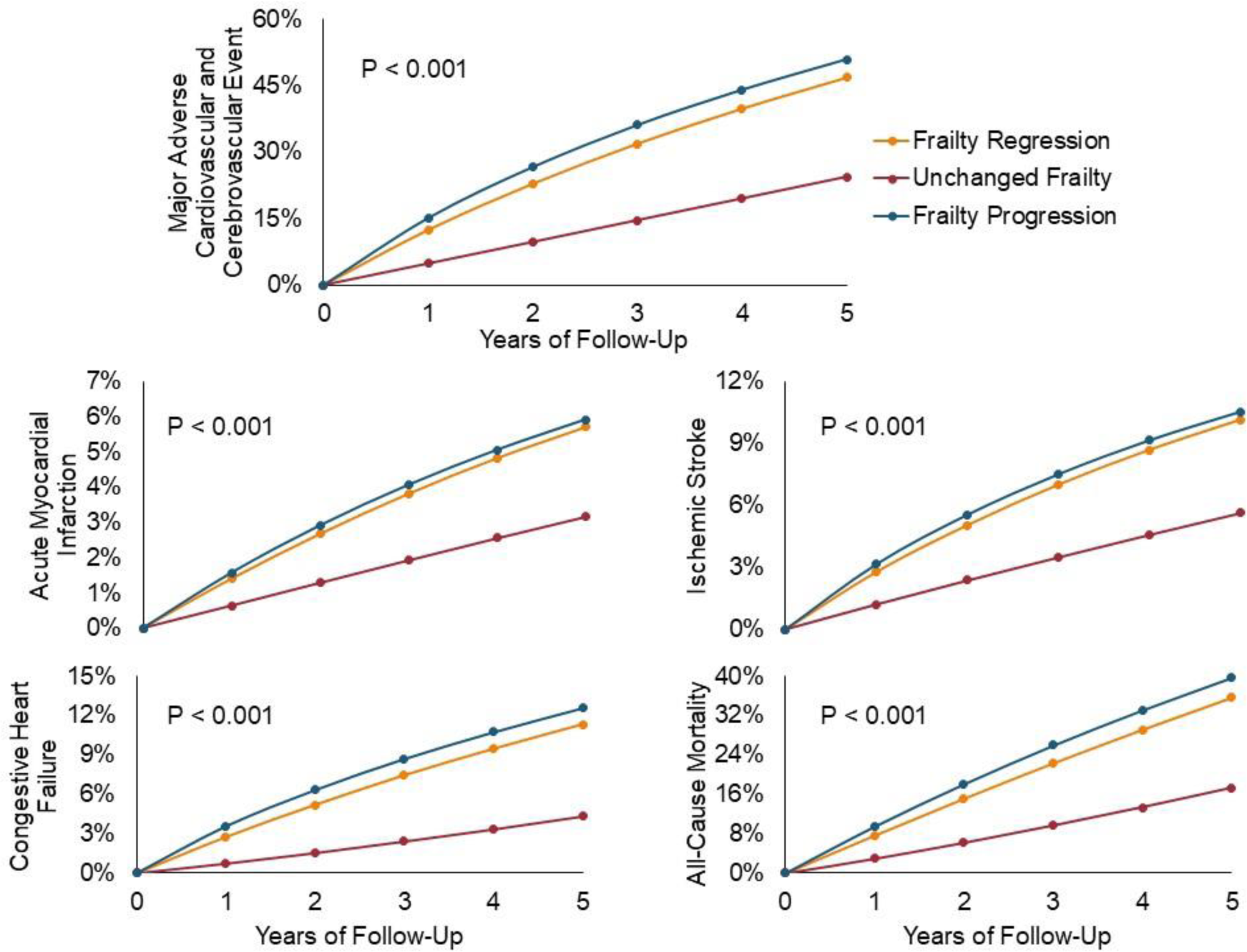
Cumulative Incidence of All-cause Mortality and Cause-specific Hospitalizations for Frailty Progression versus Regression. Cumulative incidence functions are shown for all outcomes, stratified by frailty trajectory. Frailty progressors (blue line) were defined as claims-based frailty index (CFI) slope > 0.09, frailty regressors (yellow line) were defined as CFI slope < 0.09, and those with unchanged frailty (red line) as slope = 0.09.

**Table 3.** Hazard Ratios of All-cause Mortality and Cause-specific Hospitalizations According to Frailty Trajectory. Shown are hazard ratios (HRs) and 95% confidence intervals (CIs) for all outcomes evaluated by frailty progression group. The HRs for frailty regression (claims-based frailty index slope < 0.09) and frailty progression (claims-based frailty index slope > 0.09) are compared to those with unchanged frailty (claims-based frailty index slope = 0.09). MACCE: major adverse cardiovascular or cerebrovascular event.

| Comparison | MACCE HR<br>(95% CI) | All-Cause<br>Mortality HR<br>(95% CI) | Acute<br>Myocardial<br>Infarction HR<br>(95% CI) | Congestive<br>Heart Failure<br>HR (95% CI) | Ischemic<br>Stroke HR<br>(95% CI) |
| --- | --- | --- | --- | --- | --- |
| Regressors vs.<br>Unchanged Frailty | 1.58<br>(1.57, 1.58) | 1.46<br>(1.46, 1.47) | 1.57<br>(1.56, 1.58) | 2.13<br>(2.12, 2.14) | 1.52<br>(1.52, 1.53) |
| Progressors vs.<br>Unchanged Frailty | 2.30<br>(2.30, 2.31) | 2.32<br>(2.32, 2.33) | 1.78<br>(1.77, 1.79) | 2.780<br>(2.77, 2.79) | 1.78<br>(1.77, 1.79) |

### Outcomes of Frailty Progressors by Progression Quartile

With increasing quartiles of frailty progression, there was a graded increase in all outcomes (**Figure 3**). Compared to quartile 1 of frailty progression (slowest progression group), those in quartile 4 (fastest progression group) had a significant increase in risk of MACCE (65.8% vs. 36.3%, HR 2.10, 95% CI 2.09 - 2.11), all-cause mortality (54.5% vs. 26.0%, HR 2.221, 95% CI 2.213 - 2.230), MI (52.1% vs. 24.9%, HR 1.252, 95% CI 1.24 - 1.27), HF (45.0% vs. 23.0%, HR 2.06, 95% CI 2.05 - 2.08), and ischemic stroke (49.1% vs. 23.6%, HR 1.38, 95% CI 1.37 - 1.39). Quartiles 2-3 similarly demonstrated higher risk of all outcomes compared to quartile 1 (**Table 4**).

**Figure 3.**
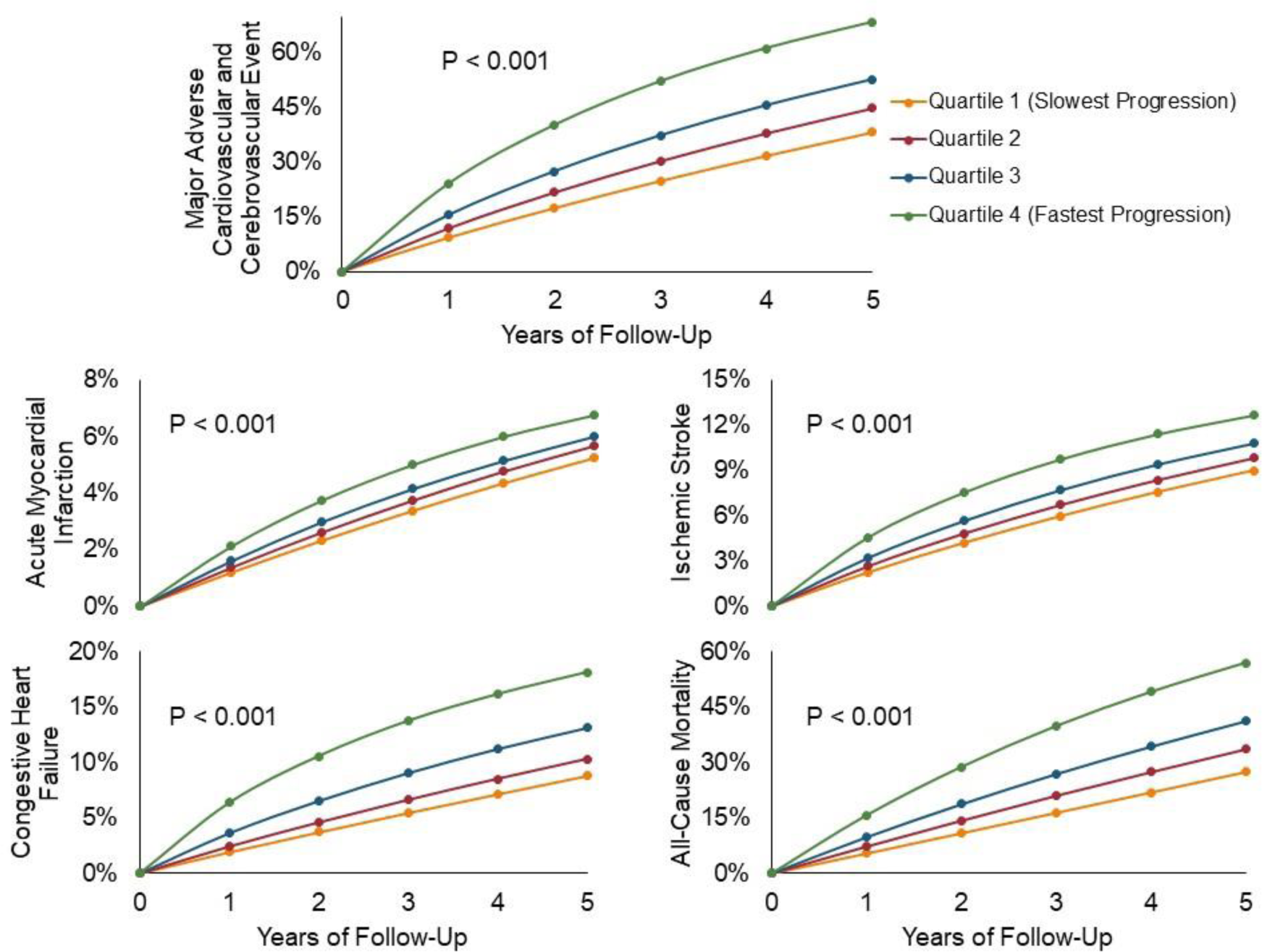
Cumulative Incidence of All-cause Mortality and Cause-specific Hospitalizations Among Frailty Progressors, Stratified by Quartile of Progression Rate. Cumulative incidence functions are shown for all outcomes, among those with frailty progression (claims-based frailty index slope > 0.09), stratified by quartile of progression rate. Numerically higher quartiles indicate faster frailty progression.

**Table 4.** Adjusted Hazard Ratios of All-cause Mortality and Cause-specific Hospitalizations by Quartile of Frailty Progression. Shown are hazard ratios (HR) and 95% confidence intervals (CI) for all outcomes evaluated according to frailty progression quartile among frailty progressors (claims-based frailty index slope > 0.09), using quartile 1 as reference. Numerically higher quartiles indicate faster frailty progression. MACCE = major adverse cardiovascular or cerebrovascular event.

| Comparison | MACCE HR (95% CI) | All-Cause Mortality HR (95% CI) | Acute Myocardial Infarction HR (95% CI) | Congestive Heart Failure HR (95% CI) | Ischemic Stroke HR (95% CI) |
| --- | --- | --- | --- | --- | --- |
| Quartile 2 vs. 1 | 1.23 (1.22, 1.23) | 1.27 (1.27, 1.28) | 1.08 (1.07, 1.09) | 1.18 (1.17, 1.19) | 1.10 (1.09, 1.11) |
| Quartile 3 vs. 1 | 1.48 (1.47, 1.48) | 1.55 (1.54, 1.55) | 1.13 (1.12, 1.14) | 1.49 (1.48, 1.50) | 1.19 (1.18, 1.20) |
| Quartile 4 vs. 1 | 2.10 (2.09, 2.11) | 2.22 (2.21, 2.23) | 1.25 (1.24, 1.27) | 2.06 (2.05, 2.08) | 1.38 (1.37, 1.39) |

Similarly, with more rapid frailty progression, individuals experienced fewer DAH in the first year of follow-up with a graded effect (**Table 5**). Compared to quartile 1, quartile 4 had a mean of 38.2 fewer DAH (308.6 ± 93.0 vs. 270.4 ± 112.3 DAH, rate ratio 0.876, 95% CI 0.874 – 0.878; p < 0.001).

**Table 5.**
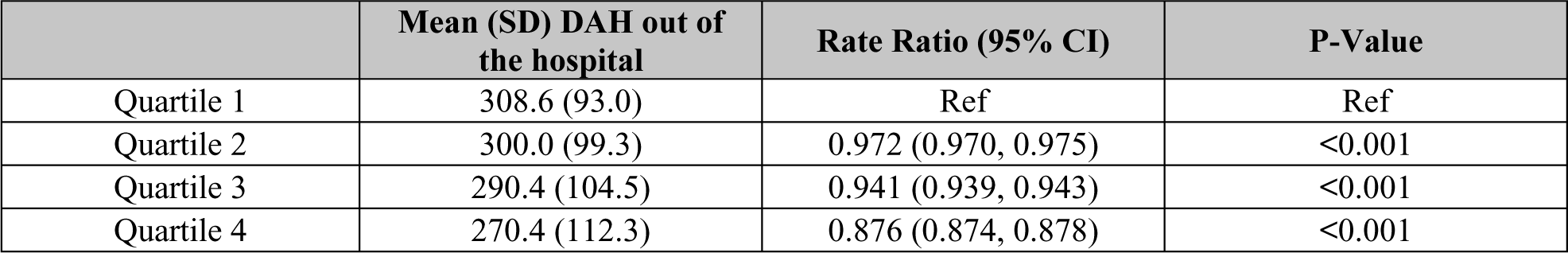
Days at home out of the hospital by FI slope progression quartile. Shown is the mean (standard deviation; SD) number of days alive at home (DAH) out of the hospital by quartile of frailty progression, among frailty progressors (claims-based frailty index slope > 0.09). Numerically higher quartiles indicate faster frailty progression. Also shown is the rate ratio and 95% confidence interval (CI) for DAH for each quartile using quartile 1 as reference (Ref).

## DISCUSSION

This study explores the independent association of frailty progression with adverse cardiovascular health outcomes in the US. In a large retrospective cohort study of Medicare beneficiaries, progression of frailty, as measured using a CFI, occurred in 20.4% and was associated with an increasing graded risk of MACCE, all-cause mortality, HF, MI, and ischemic stroke, independent of baseline levels of frailty. With increasing frailty progression, individuals spent fewer DAH with those at the fastest rates of progression spending 38 fewer days at home than those with the slowest rates of progression. Our results indicate that both the severity of frailty and the degrees of frailty progression may help to identify those at high risk for adverse outcomes and increased health services utilization. Serial frailty testing to guide decision-making and to identify high-risk individuals for cardiovascular interventions represents an important area for further investigation.

Frailty, regardless how it is measured, has been shown to be an independent risk factor for adverse health outcomes after cardiac and non-cardiac procedures and has been previously used to risk stratify patients for invasive and high-risk interventions.^10,35–38^ As a corollary to this, there have been strategies such as pre-rehabilitation, entailing physical therapy with a goal of improving one’s functional status prior to a procedure, and geriatric co-management, that have been suggested to attenuate the effect of frailty on risk.^21,22,39,40^ While the impact of pre-rehabilitation efforts has been mixed, intrinsic to this premise is the notion that frailty is at least partially mutable.^22,41^

While these strategies are predicated on the possibility of frailty improvement, few studies have quantitatively evaluated whether frailty changes over time, the expected rates of change, and the independent association of these changes with adverse health outcomes. In a prior study on the US Veteran Health Administration database of predominantly (99.2%) male subjects, cardiovascular disease was identified as the leading cause of death among highly frail individuals at the end of life.^17^ A study of 3 prospective cohorts identified frailty progression using the Rockwood frailty conception (in which frailty is conceived as an accumulation of deficits over time) as being associated with increased risk of incident cardiovascular disease.^20^ Whether these results apply to Medicare beneficiaries, particularly using the Fried conception of frailty (in which frailty is conceived a clinical syndrome), and whether frailty progression is independently associated with DAH has not been previously addressed to our knowledge.

One prior study of 14,000 community-dwelling adults in Europe >55 years old evaluated the natural history of frailty,^42^ and found 22.1% of participants had progression in frailty, 61.8% had unchanged frailty, and 16.1% had frailty regression. Those at risk of worsening frailty progression included women, older individuals, and those with lower education levels. Similarly, in this large, predominantly White US Medicare beneficiary population, we found that 20.4% had progression of frailty, 13.6% had regression, and 66.1% had no change in frailty over a 5-year period, however no sex-differences were identified. In a prior study enrolling community-dwelling individuals in Spain,^43^ increasing frailty or consistently high frailty levels were associated with an increased risk for all-cause mortality, with those developing frailty from non-frailty having a 2-fold increased risk of death. We similarly found a 59% higher risk of all-cause mortality in those with frailty progression compared to the unchanged frailty group.

The current study of 26.4 million Medicare beneficiaries builds upon this prior literature by suggesting that those with frailty progression have a graded increase in all non-death outcomes evaluated, including those in the fastest frailty progression group having 38 fewer DAH at 1-year. This is an outcome of particular relevance to frail individuals when making end-of-life decisions about care.^44,45^ While more granular metrics of frailty derived from in-person measurement may be necessary for counseling patients about expected outcomes, our study nevertheless suggests an independent relationship of frailty progression with non-death health outcomes of relevance. Future investigation is needed to evaluate the role of prospectively assessing frailty progression using in-person frailty metrics in guiding shared decision-making efforts and identifying high-risk individuals for cardiovascular interventions. Furthermore, identifying the individual, modifiable contributors to frailty progression using detailed clinical and molecular phenotyping will be important in understanding the pathophysiology of frailty progression and suggesting targets for possible future interventions to slow or halt progression.

Our findings notably incorporate adjustment for levels of baseline frailty. As frailty is frequently used to risk stratify patients for high-risk interventions such as aortic valve replacement,^10,12,46,47^ mitral valve procedures,^36,48,49^ and left atrial appendage closure,^37,38^ our results suggest a possible additional role for measures of frailty progression, derived from serial frailty measurements, to guide risk conversations. Given the higher a risk of adverse outcomes in those with frailty progression is independent of baseline frailty, these data may suggest a role for frailty trajectory assessment for those without frailty to define a higher-risk cohort. Such measurements could be routinely incorporated into practice, similar to the use of hemoglobin A1c to monitor one’s diabetes status and could even be derived automatically from variables in the electronic health record to streamline calculation and facilitate shared decision making.^50^

Of note, we paradoxically found that individuals without a change in frailty had improved outcomes compared to those with frailty regression. This may be partially explained by so-called floor effects in the calculation of the CFI. In order to have regression in frailty as defined by CFI, one must start from a state of greater frailty severity, which was evidenced by the greater number of comorbidities in those with frailty regression and worse overall outcomes. Nevertheless, it is of note that those with frailty regression had overall better outcomes than those with frailty progression. While it is possible that those with frailty regression are different than those with frailty progression in substantive, unmeasured ways that influence their risks of experiencing outcomes, it is also plausible that efforts to slow or reverse one’s frailty progression may have associated health benefits. Accordingly, future research should further examine the independent influence of frailty regression on health outcomes.

Our study should be interpreted in the context of a few limitations. First, as a study of Medicare FFS beneficiaries, these findings may not be generalizable to patients < 65 years old, those with commercial insurance, or those with Medicare Advantage plans. Second, though we chose a CFI that most closely approximates the Fried index,^6^ a measure of phenotypic frailty, in order to evaluate frailty as a syndrome, these findings may not apply to other frailty constructs (e.g. the Rockwood definition^7^) or other instruments and should be validated in other settings. Third, given the retrospective nature of the current study, causality cannot be inferred using the current methods. Fourth, as we evaluated all Medicare FFS beneficiaries over the study period, it is possible that the influence of frailty trajectory may differ in disease subsets, which should be evaluated in future research efforts. Finally, our study was unable to evaluate or adjust for known influences of frailty trajectories such as socioeconomic status, home support, and education.

## CONCLUSIONS

Among Medicare beneficiaries, the degrees of frailty progression, independent of baseline frailty, was associated with fewer DAH and increased incidence of MACCE and adverse cardiovascular outcomes. Our results indicate that frailty progression may improve upon frailty status alone in identifying those at high risk for adverse outcomes and increased health services utilization.

## Data Availability

Data used in this study are from the United States Centers for Medicare and Medicaid Services (CMS) Medicare Provider Analysis and Review (MedPAR) database from January 1, 2003 through December 31, 2020. The data used in this analysis are not available for public sharing due to data use agreements with Medicare (RSCH-2018-52411).

